# Olfactory decline in aging: longitudinal trajectories and associations with cognitive decline and postmortem neuropathology

**DOI:** 10.64898/2026.04.27.26351817

**Authors:** Cecilia Tremblay, Parichita Choudhury, Erika Driver-Dunckley, Zaki Alasmar, Geidy E Serrano, Holly A. Shill, Shyamal Mehta, Andrew Ho, Seyed-Mohammad Fereshtehnejad, David Shprecher, Ileana Lorenzini, Christine M. Belden, Alireza Atri, Charles H Adler, Thomas G Beach, Mahsa Dadar, Yashar Zeighami

**Author notes:** CORRESPONDENCE: Cecilia Tremblay, Cerebral Imaging Centre, Douglas Mental Health University Institute, Montréal, Québec, Canada, H4H 1R3, Mahsa Dadar, Cerebral Imaging Centre, Douglas Mental Health University Institute, Montréal, Québec, Canada, H4H 1R3, Yashar Zeighami, Cerebral Imaging Centre, Douglas Mental Health University Institute, Montréal, Québec, Canada, H4H 1R3. co-senior authors (Mahsa Dadar, Yashar Zeighami).

## Abstract

**Importance:** Decline in olfactory function may be used as a predictor of cognitive decline, to enhance early detection models, improve risk stratification, and enable early intervention.

**Objective:** To assess the longitudinal association between olfactory decline, cognitive decline, and postmortem neuropathology.

**Design, setting and participants:** Retrospective longitudinal analysis with clinicopathological correlations of a prospective population-based cohort study using data from the Arizona Study of Aging and Neurodegenerative Disorders (AZSAND) and its Brain and Body Donation Program. Participants included cognitively unimpaired individuals without parkinsonism that converted to mild cognitive impairment (MCI) and/or dementia or remained cognitively stable.

**Main Outcomes and Measures:** longitudinal change in olfaction, neuropsychiatric symptoms, motor function and memory, conversion to MCI/dementia, postmortem neuropathology

**Results:** Over a mean follow-up period of 7.7 ± 5.4 years, out of 922 participants who were cognitively unimpaired at the first cognitive conference, 643 remained cognitively unimpaired, 279 converted to MCI, and 82 developed dementia. Of these, 633 individuals had at least 2 olfactory tests.

Converters showed reduced olfactory function (t=-12.6, p <0.0001), faster progression in neuropsychiatric symptom burden (t=3.42, p < 0.001), and faster decline in memory (t= -7.33, p <0.0001) prior to conversion while no significant differences were observed in motor scores between converters and non-converters. Using ROC analysis, olfactory decline, increased neuropsychiatric symptom burden, as well as motor and memory decline predicted conversion to MCI with a consistent accuracy of ∼ 70% up to 5 years before conversion, while UPSIT alone had an accuracy of ∼ 60%. Longitudinal decline in olfaction was associated with a higher burden of a-synuclein (t= -8.21, p <0.0005), tau tangle (t= -2.66, p < 0.01) and amyloid plaque burden (t= -2.85, p < 0.005) and a faster decline over time was associated with a higher burden of tau (t=5.66, p<0.0001).

**Conclusions and Relevance:** A reduction in olfactory identification ability is observed up to a decade prior to conversion to MCI and is associated with underlying burden of neuropathology markers, underscoring the value of incorporating olfactory testing in cognitively unimpaired individuals to identify those at-risk of future cognitive decline.

## Introduction

Longitudinal studies examining olfactory function changes in aging are critically needed to assess within-person decline over time and to distinguish normal age-related decline from that associated with neurodegenerative disease. Decline in olfactory function can predict mild cognitive impairment (MCI) and dementia^1–6^ and is associated with faster progression from MCI to dementia.^5,7,8^ Faster olfactory decline is associated with atrophy of olfactory related regions.^9,10^ Early Alzheimer’s disease (AD) and Lewy body diseases (LBD) affect central olfactory processing regions and previous findings report cross-sectional associations between olfactory decline and increased burden of neurofibrillary tangles (NFT) and alpha-synuclein (aSyn) pathologies particularly.^11,12^ Yet, the pathophysiological mechanisms underlying disease-related olfactory impairment remain unclear, particularly in early stages.^13,14^ Studies with neuropathological confirmation are sparse and critically needed to better characterize the underlying mechanisms driving olfactory decline in healthy aging and neurodegenerative diseases.

The present study evaluates longitudinal olfactory trajectories in aging among individuals who remained cognitively normal or transitioned to MCI and/or dementia. We additionally assessed longitudinal trajectories of neuropsychiatric symptom burden, motor, and memory decline in the same individuals to determine whether they can be combined with olfactory decline to predict conversion to MCI. We further investigated the association between longitudinal changes in olfaction and postmortem neuropathology markers of AD and frequent comorbidities, including tau, amyloid, aSyn and vascular pathologies. We hypothesized that 1) early olfactory decline predicts cognitive decline and transition to MCI and dementia; 2) longitudinal olfactory decline is associated with decline in motor function and neuropsychiatric burden and 3) olfactory decline is associated with increased burden of neuropathology markers.

## Methods

### Study design and participants

Participants were included from the Arizona Study of Aging and Neurodegenerative Disorders (AZSAND) and its Brain and Body Donation Program, a clinicopathological study of aging conducted at Banner Sun Health Research Institute (BSHRI).^15^ Participants were elderly volunteers including independent living, cognitively and movement-unimpaired individuals as well as those living with dementia and parkinsonism. All participants provided written informed consent, and study procedures were approved by BSHRI Institutional Review Boards.

### Clinical assessments

Participants underwent comprehensive clinical assessments including annual standardized test batteries, consisting of general neurological, neuropsychological, cognitive, and movement disorders components administered by neuropsychologists and subspecialty cognitive/behavioral and movement disorders certified neurologists. Participants received a consensus cognitive diagnosis following their yearly evaluations based on review of all available assessments by a cognitive neurologist and neuropsychologist. Standardized movement examinations, including the Unified Parkinson’s Disease Rating Scale (UPDRS) were also performed annually.^16^ A final clinical diagnosis was established after death, but prior to neuropathological examination, at a consensus conference in which all clinical assessments and medical records were reviewed. Finally, clinicopathological diagnoses were assigned after death taking into consideration pathological and clinical findings.

For this study, all individuals who were cognitively normal and without parkinsonism at enrollment with longitudinal assessments available were included. Annual cognitive consensus conference diagnosis, or final conference consensus diagnosis when available, was used to assign a clinical diagnosis of cognitively normal, MCI, or dementia. Individuals with possible/probable PD, DLB or atypical parkinsonism at any movement assessment visit were excluded.

### Assessment of olfaction and neuropsychiatric burden

Olfactory function was measured every three years, using the 40-item University of Pennsylvania Smell Identification Test (UPSIT).^17^ Neuropsychiatric symptoms were evaluated yearly through the Neuropsychiatric Inventory-Questionnaire (NPI-Q) recording frequency and severity of delusions, hallucinations, agitation or aggression, depression or dysphoria, anxiety, elation or euphoria, disinhibition, irritability or lability, motor disturbance, nighttime behaviors, appetite and eating.^18^

### Neuropathological assessments

Comprehensive postmortem neuropathological examination was performed,^15^ including AD Braak NFT stages^19^, CERAD neuritic and diffuse plaque density scores, Thal amyloid phase, and aSyn pathology stage according to the Unified Staging System for Lewy Body Disorders (USSLB).^20^ Data also included regional and summary cortical density measures for tau NFT load and amyloid plaque load (summary of 0–3 scores in each of 5 regions: frontal, parietal and temporal association cortices, hippocampus CA1, and entorhinal/transentorhinal areas). Cerebrovascular pathology included the presence and number of microinfarcts, density scores for cerebral amyloid angiopathy (CAA), and cerebral white matter rarefaction (CWMR).^21^

### Statistical analysis

Baseline characteristics of individuals that stayed cognitively normal (non-converters) or progressed to MCI/dementia (converters) were compared using *t-*tests for continuous or ***χ***^2^ tests for categorical variables. Mixed-effects models were used to compare longitudinal trajectories of olfaction, neuropsychiatric symptoms, movement impairment, and memory (Rey Auditory Verbal Learning Test (AVLT^22^)) between converters and non-converters for individuals with a minimum of two assessments available. For converters, the last cognitive conference at which an individual was cognitively unimpaired was defined as the conversion point (Year 0) and trajectory slopes were compared between before and after conversion. For non-converters, Year 0 was defined by <u>age-matching</u> them to the converters at the conversion point. Covariates included age at the time of clinical evaluation, time from first evaluation, and sex.

To assess the associations between clinical measures, the slope of change in each measure was estimated per subject for the 10-year pre-conversion period (**supplementary table 1**). Correlation analyses then examined whether rates of change were correlated across measures. Further, receiver operating characteristic (ROC) curves were computed to assess the combined ability of clinical measures (UPSIT, NPI-Q, UPDRS-III, AVLT) to predict conversion to MCI from 1-5 years before conversion, with the 5-year limit determined by sample size limitations across overlapping measures.

For individuals with post-mortem neuropathology assessments, longitudinal and cross-sectional (with last assessment) associations between olfactory decline and neuropathologies were examined. Associations were explored with total pathology burden including tau tangle burden, amyloid plaque burden, aSyn burden, and severity of CWMR and CAA, presence and number of microinfarcts, as well as disease stages including Braak NFT stage, Thal amyloid phase, CERAD neuritic plaque score, and aSyn USSLB stage. To further assess the independent role of neuropathologies, total burden of tau, amyloid and aSyn were included in the same model. Analyses were further repeated in cognitively unimpaired individuals to assess olfactory impairment associations preceding MCI.

## Results

### Study participants

A total of 922 participants were cognitively unimpaired at the first cognitive conference, with a total of 5879 longitudinal cognitive conferences available over a mean period of 7.7 ± 5.4 years. Of these, 643 participants remained cognitively unimpaired (follow up = 6.8 ± 5.2), 361 individuals converted to MCI/dementia (follow up = 9.7 ± 5.5; pre-conversion follow up = 5.9 ± 5.4). Participant numbers and follow up durations differed across clinical tests. **Tables 1-2** summarize baseline characteristics of the full sample and the subset with neuropathological data available.

**Table 1:** Demographics and general characteristics of included cases at <u>baseline</u> per conversion status for each clinical variable.

|  | Non-converters | Converters | P value |
| --- | --- | --- | --- |
| **UPSIT** |  |  |  |
| N (longitudinal visits) | 421 (1253) | 212 (691) |  |
| Age at baseline | 74.7 ± 7.5 | 78.1 ± 7.3 | P < 0.001 |
| Sex, Male % | 124, 29% | 77, 36% | P ~ 0.1 |
| mean UPSIT* | 32 ± 5.4 | 29.5 ± 6.6 | P < 0.001 |
| **NPI-Q** |  |  |  |
| N (longitudinal visits) | 534 (3146) | 228 (1749) |  |
| Age | 76.6 ± 7.6 | 80.3 ± 7.2 | P < 0.001 |
| Sex, Male % | 158, 29% | 88, 38.5% | P < 0.05 |
| mean NPI-Q* | 0.92 ± 1.5 | 1.44 ± 2.2 | P < 0.001 |
| **UPDRS - III** |  |  |  |
| N (longitudinal visits) | 628 (4124) | 266 (2290) |  |
| Age | 75.6 ± 7.7 | 77.8 ± 7.5 | P < 0.001 |
| Sex, Male % | 193, 30% | 105, 39% | P < 0.05 |
| Mean UPDRS-III* | 2.5 ± 3.3 | 3.1 ± 4.6 | P ~ 0.27 |
| **AVLT** |  |  |  |
| N (longitudinal visits) | 637 (4111) | 277 (2264) |  |
| Age | 75.7 ± 7.8 | 77.1 ± 7.6 | P < 0.001 |
| Sex, Male % | 199, 31% | 112, 40% | P < 0.01 |
| Mean AVLT* | 45.0 ± 1.7 | 41.6 ± 9.1 | P < 0.01 |

\* p value is calculated after considering age and sex as well as their interaction as covariates.
Abbreviations: UPSIT, University of Pennsylvania Smell Identification Test; NPI-Q, Neuropsychiatric Inventory-Questionnaire; UPDRS-III, Unified Parkinson's Disease Rating Scale motor score part 3; AVLT, Auditory Verbal Learning Test

**Table 2:** Demographics and general characteristics of cases with neuropathological evaluation, for everyone with available UPSIT assessments and for cognitively unimpaired controls.

|  | Whole cohort with UPSIT | Controls with UPSIT |
| --- | --- | --- |
| N (cross-sectional) | 485 | 136 |
| N (longitudinal visits) | 250 (966) | 72 (297) |
| Age at death | 87.9 ± 7.7 | 87.6 ± 7.2 |
| Sex, Male % | 256, 52.8% | 70, 51.5% |
| last UPSIT | 22.8 ± 8.2 | 27.3 ± 6.7 |
| Number (%) of cases per Braak NFT stages (I/II/III/IV/V/VI ) | 9 (2.3)/ 24 (4.9)/ 71 (14.6)/ 228 (47.0) / 104 (21.4)/ 49 (10.1) | 7 (5.1)/ 16 (11.7)/ 37 (27.2)/ 75 (55.1)/ 1(0.7) /0 |
| Thal amyloid phase, number (%) (0,1,2,3,4,5) | 100 (20.6)/ 49 (10.1)/ 41 (8.4)/ 105 (21.6)/ 49 (10.1)/ 79 (16.3) | 49 (36.0)/ 16 (3.3)/ 19 (14.0)/ 27 (19.8)/ 7 (5.1)/ 7 (5.1) |
| CERAD neuritic plaque, number (%) (zero,sparse, moderate, frequent) | 117 (24.1)/ 48 (9.9)/ 41 (8.4)/ 278 (57.3) | 51 (37.5)/ 24 (17.6)/ 24 (17.6)/ 37 (27.2) |
| USSLB stages (0, I, IIa, 2b, III,IV) | 318 (65.6)/ 15 (3.1)/ 39 (8.0)/ 34 (7.0)/ 42 (8.7)/ 37 (7.6) | 108 (79.4)/ 3 (2.2)/ 12 (8.8)/ 4 (2.9)/ 8 (5.9)/ 1 (0.7) |
| Total tangle burden | 8.0 ± 3.7 | 5.4 ± 2.4 |
| Total plaque burden | 8.1 ± 6.0 | 5.1 ± 5.6 |
| Total aSyn burden | 5.2 ± 9.7 | 2.5 ± 6.1 |
| Total CWMR score | 4.2 ± 3.2 | 2.7 ± 2.4 |
| Total CAA score | 3.1 ± 3.5 | 2.3 ± 3.2 |
| Number of cases with infarcts, % | 231, 48% | 43, 32% |

Abbreviations: UPSIT, University of Pennsylvania Smell Identification Test; USSLB, Unified Staging System for Lewy Body Disorders; aSyn, alpha-synuclein pathology; CWMR, cerebral white matter rarefaction ; CAA, Cerebral amyloid angiopathy

### Clinical measures distinguish converters and non-converters years before conversion

Olfactory UPSIT scores were significantly lower by 1.31 in the converters at the time of conversion (t= -2.13, p<0.05). Both groups showed UPSIT declines over time at a rate of 0.60 per year (t= -12.6, p<0.0001) with no statistically significant group differences suggesting a persistent difference between groups during the decade prior to conversion (Figure 1-A). The converter group showed steeper olfactory decline after conversion, with 0.58 units per year increase in rate of decline (t= -2.71, p<0.005) (Figure 1-B).

**Figure 1:**
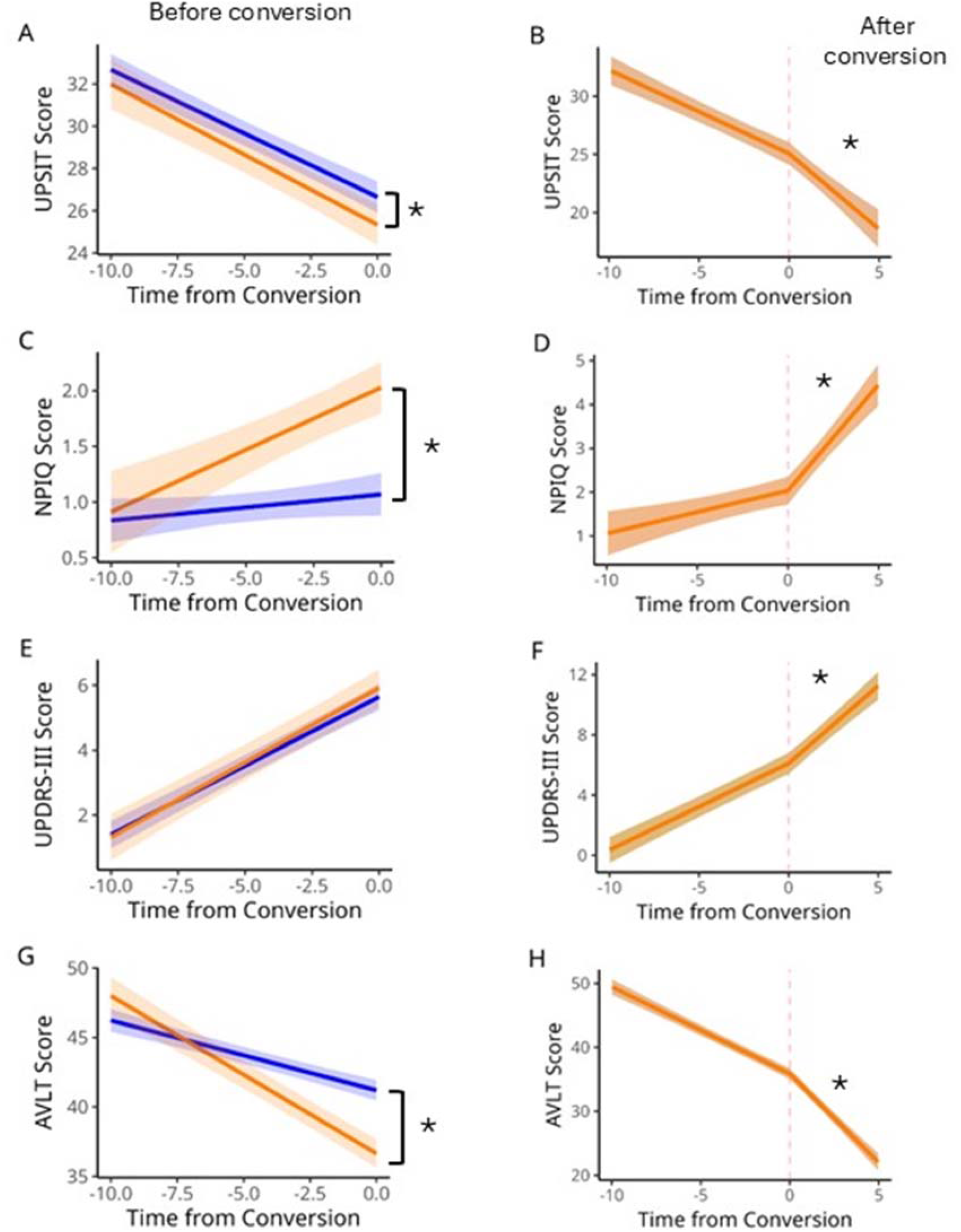
Trajectories of olfactory function (UPSIT), neuropsychiatric symptom burden (NPI-Q), motor function (UPDRS-III) and memory (AVLT) before conversion (left panel) for converters (orange) and non-converters (blue). The right panel shows both the pre and post conversion for the converter group. Time 0 represents the conversion point to MCI. In converters, significant decline is observed for all domains after conversion.

Neuropsychiatric symptom burden (NPI-Q) was significantly higher in converters at the time of conversion (b = 0.96, t=6.25, p< 0.0001). Moreover, converters showed faster progression of overall neuropsychiatric symptoms prior to conversion (b= 0.09/year, t=3.42, p<0.001) while this was not significant for non-converters (b = 0.02/year; t= 1.56, p>0.05) (Figure 1 - C). The converter’s progression further accelerated following conversion (b = 0.039 increase per year, t=5.4, p<0.0001) (Figure 1-D).

While individuals with PD, DLB and atypical parkinsonism were excluded, there was an age-related increase in motor signs (UPDRS-III) (b=0.42 per year, t=16.7, p<0.0001), with no significant main effect or slope differences between the two groups (Figure 1-E). Converters showed increased motor impairment post conversion (b= 0.45 per year, t=4.14, p<0.0001) (Figure 1-F).

Memory tests (total AVLT) showed significant group differences at the time of conversion (b= -4.55, t=-7.01, p < 0.0001), age-related decrease over time (b= -0.5, t=-9.9, p<0.0001), as well as an interaction with conversion status, suggesting a further -0.63/year deterioration in converters in the decade prior to conversion (b= -0.63, t = -7.33, p <0.0001) (Figure 1-G). This deterioration was accelerated after conversion by -1.42/year (b= -1.42, t = -7.7, p <0.0001) (Figure 1-H).

### Olfactory decline is associated with increased neuropsychiatric burden, decline in motor function and memory and improve conversion to MCI prediction

UPSIT decline rate (slope of change) in the 10 years prior to conversion was associated with that of UPDRS (r = -0.21, p<0.001), NPI-Q (r = -0.09, p=0.04) and AVLT (r = 0.18, p<0.001) in non-converters.

ROC analysis evaluated the ability of UPSIT, NPI-Q, UPDRS-III, AVLT, and their combination to predict conversion to MCI from 1 year to 5 years before conversion (with 1-year intervals). The full model consistently performed at ∼ 70% accuracy (Figure 2.A). As expected, the AVLT predictive ability was best at 1 year prior to conversion (AUC= 0.67). However, its predictive ability progressively weakened with greater pre-conversion intervals (0.59 and 0.54 at 2 and 4 years, respectively), while UPSIT and NPI-Q had consistent AUCs of ∼ 0.6 up to 4 years before conversion (Figure 2.B).

**Figure 2:**
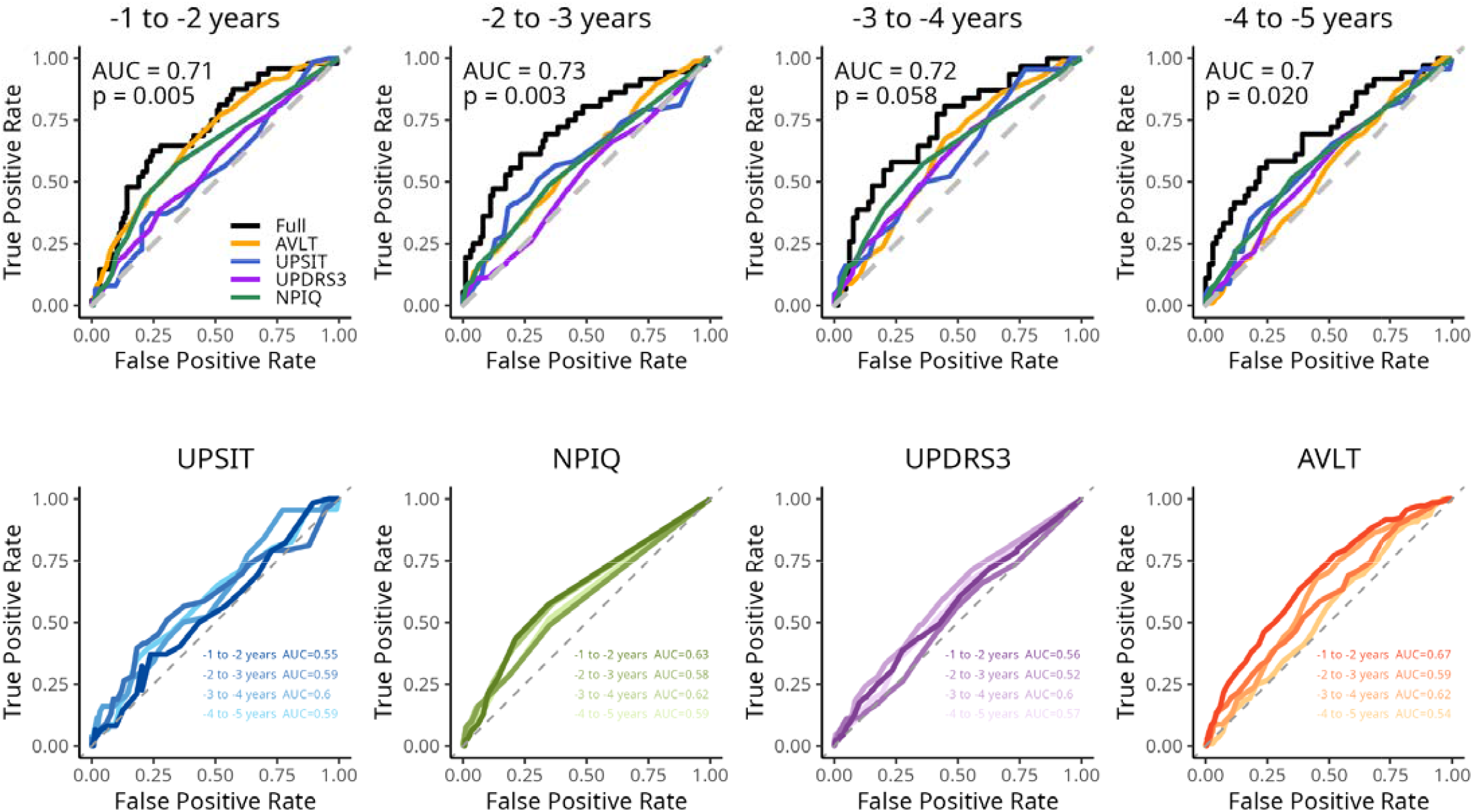
Receiver Operating Characteristic (ROC) curves combining olfactory identification, neuropsychiatric symptom burden, motor functioning and memory to predict conversion to MCI from 1 to 5 years before conversion with 1 year interval (A) and prediction performance separately for each measure from 1 to 5 years pre-conversion to MCI (B). The full model consistently performed at ∼ 70% accuracy while UPSIT alone performed at ∼ 60% accuracy to predict MCI.

### Olfactory decline is associated with neuropathology burden and disease stages

When investigating associations between the latest UPSIT test and pathology burden, controlling for sex, age at the last visit as well as time to death, the largest effect size was observed with total aSyn burden (b= -4.07, t= -12.43, p< 0.0001; adjusted R^2^ = 0.26) followed by total tau tangle burden (b= - 2.98, t= -8.42, p<0.0001; adjusted R^2^= 0.14) and total amyloid plaque burden (b= -2.38, t= -6.66, p< 0.0001; adjusted R^2^= 0.10) (Figure 3.A). When the three pathologies were considered simultaneously, the greatest portion of variability was explained by total aSyn burden (b= -3.70, t= -11.8, p<0.0001) and total tangle burden (b= -2.24, t= -5.95, p< 0.0001).

**Figure 3:**
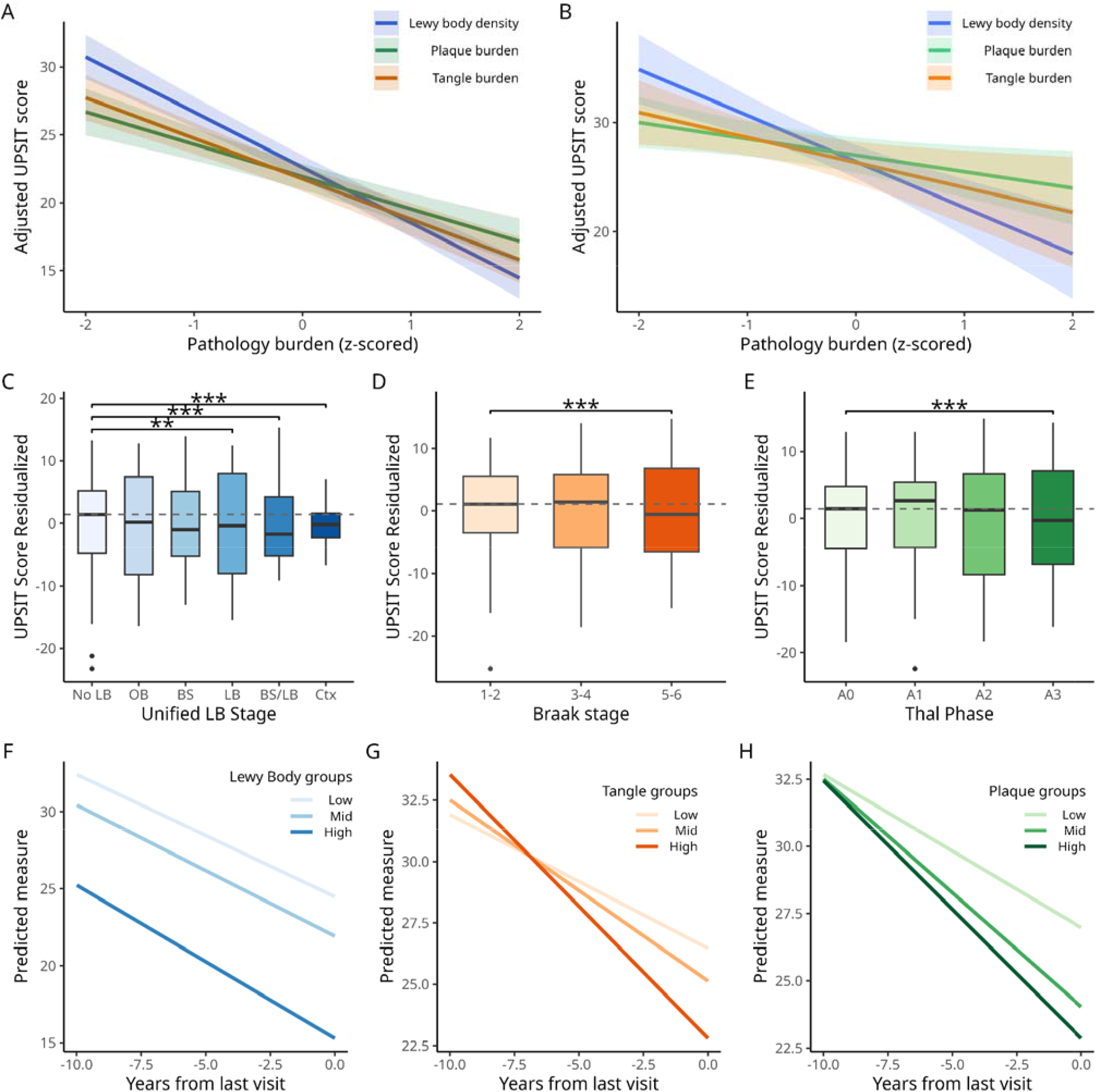
**R**elationship between olfactory function and neuropathology burden looking at cross-sectional relationship with last UPSIT test in everyone (A) and in cognitively unimpaired individuals (B) for total burden of pathologies and for disease stages (C-E). Longitudinal associations between olfactory decline for total burden of a-synuclein (F), neurofibrillary tangles (H) and amyloid plaque (H) pathologies with trajectories represented as tertiles.

Modeling longitudinal olfactory decline as a function of postmortem pathology burden in the whole sample, a higher burden of aSyn (b= -3.12, t= -8.21, p<0.0005) was associated with lower olfactory function while there was no significant interaction with time. There was both a main effect of total tangle burden (b= -1.15, t= -2.66, p< 0.01) and its interaction with time (b= -0.92, t=5.66, p<0.0001), suggesting faster olfactory decline in participants with higher tau pathology. Similarly, we found both a main effect of total amyloid burden (b= -1.24, t= -2.85, p< 0.005) and interaction with time (b= -0.50, t=3.05, p<0.005) (Figure 3.F-H). When considering pathologies simultaneously, the main effect of aSyn pathology remained significant (b=-3.07, t=-8.237, p< 0.0001) with no effect of tau or amyloid burden, while only tau burden showed a significant interaction on longitudinal changes in olfactory performance (b= -0.88, t= -4.67, p<0.0001).

Including only cognitively unimpaired controls, associations with last UPSIT were also found for aSyn burden (b=-2.62, t= -4.99, p< 0.0001; adjusted R^2^= 0.21), tangle burden (b= -1.45, t= -2.45, p< 0.02; adjusted R^2^= 0.10) and plaque burden (b= -1.39, t= -2.46, p< 0.02; adjusted R^2^= 0.10) (Figure 3.B). Similarly, when the three pathologies were considered simultaneously, only aSyn was significant (b= - 2.44, t= -4.70, p< 0.001; adjusted R^2^= 0.25). To assess olfactory identification trajectories preceding conversion, we further examined longitudinal associations within controls, and found a significant association only with aSyn burden (b= -2.33, t= -3.71, p< 0.0005) while no significant effect of total tau and amyloid burden or any interaction with time.

When examining differences in disease stages, associations with the last UPSIT test showed a significant difference between early Braak NFT stages I-II and late stages V-VI (b= -7.9, t=-5.26, p<0.0001; Figure 3-D). Using longitudinal assessment of the olfactory system, we found a significant longitudinal interaction for both groups III-IV (b= -0.54 score per year, t = -2.6, p < 0.01) and V-IV (b= -1.03 score per year, t = -4.68, p < 0.0001) in comparison to Braak stages I-II. For USSLB aSyn stages, significant differences were found between stage IIb Limbic Predominant (b= -4.05, t= -3.17, p<0.002), stage III Brainstem/Limbic (b= -9.81, t= -8.51, p<0.0001), as well as stage IV Neocortical (b= -12.87, t= -10.48, p<0.0001) compared to individuals without pathology (Figure 3-C). For Thal phase, higher Thal phase 4-5 showed a significantly lower olfactory function (b=-5.7, t= -5.53, p<0.0001) compared to the no pathology group (Thal = 0) (Figure 3.E). There was also a significant longitudinal interaction with time (b= -0.44 score per year, t = -4.04, p < 0.0001). Similarly, for CERAD neuritic plaques, the frequent pathology group showed a significant difference (b= -4.46, t= - 5.08, p <0.0001) as well as a significant longitudinal interaction with time (b= -0.23 score per year, t = -2.46, p < 0.02).

Finally, we found an association between UPSIT and CAA (b= -0.49, t= -4.78, p< 0.0001) and CWMR (b= -0.41, t= -4.14, p< 0.0001) while no association was found with the presence of infarcts (b= -0.67, t= -0.56, p= 0.57). Similarly, longitudinal decline in olfaction was associated with CAA (b= -0.52, t= - 2.95, p<0.005) and marginally associated with CWMR (b= -0.32, t= -1.94, p∼ 0.052).

## Discussion

In this longitudinal study of olfactory function, we report significant age-related declines over time. Odor identification ability was significantly lower in individuals that converted to MCI compared to those that remained cognitively unimpaired, for up to 10 years before conversion, adjusting for age and sex. Olfactory decline was associated with an increased burden of neuropsychiatric symptoms and decline in motor and memory functions and predicted conversion to MCI with a moderate accuracy. Finally, decline in olfactory function was associated with higher burden of neuropathology markers of AD and LBD.

Diminished olfactory function was associated with incident MCI. This is in line with previous reports from the Rush Memory and Aging Project.^2,7^ Using the 12-item BSIT test, they reported significantly lower olfactory identification performance up to five years preceding MCI diagnosis as well as faster olfactory decline in the individuals converting to MCI and dementia. While we also reported lower olfactory function, we did not find a steeper decline in converters. This might be due to differences in study design, (1) we extended this work using the full 40-item olfactory identification test and (2) matched individuals for age at the time of conversion, given the well-documented effect of age on olfactory decline^23^, whereas previous reports accounted for age differences only through linear statistical adjustment. We report significantly lower olfactory function for up to 10 years before conversion to MCI. This finding has important clinical implications for early detection especially as olfactory testing is non-invasive, relatively easy to administer, and inexpensive. This underscores the value of incorporating olfactory testing, even in cognitively unimpaired aging individuals as they may be unaware of hyposmia^24,25^, to identify at-risk population and enable early therapeutic strategies.

Olfactory decline was associated with increased neuropsychiatric symptom burden, motor impairment, and decline in memory, as suggested by previous work.^26–29^ Significant declines in these domains were observed in the pre-conversion period and significant group differences in neuropsychiatric symptoms and memory were observed at baseline and the time of conversion, suggesting that conversion to MCI is a process reflected across several domains, and that subthreshold decline in memory and other domains are also measurable.^30–32^ ROC curves showed an accuracy of ∼70% in predicting conversion to MCI, 1-5 years before conversion. While memory was the main contributor one year prior to the conversion but diminishing with time, NPI-Q and UPSIT (60% accuracy) performance remained consistent up to 5 years before conversion. This accuracy is similar to previous work reporting AUC values ranging from 0.6 to 0.76 for discriminating between cognitively unimpaired and MCI combining olfactory and memory measures between 2-4 years before conversion.^5,33^ This result highlights that a combination of clinical measures including olfactory testing is invaluable for identifying individuals who should undergo more expansive/invasive diagnostic testing to detect underlying AD neuropathology or for recruitment for prevention trials. Overall, this suggests that conversion to MCI reflects multidomain clinical changes, and integrating longitudinal clinical measures considering several domains, with emerging fluid and neuroimaging biomarkers, may further improve early detection and characterization of the conversion process.^34,35^

Longitudinal decline in olfaction was associated with a higher burden of aSyn, tau tangles, amyloid plaques, CWMR and CAA. An independent effect of aSyn was observed and faster olfactory decline was associated with higher burden of tau tangle pathology. Potential mechanisms underlying this association could include early accumulation of these markers in the olfactory bulb and other olfactory regions including the entorhinal cortex, hippocampus, amygdala, and piriform cortex, which are known to be involved in the early pathophysiology of AD and LBD.^11,12,36^ In cognitively normal controls, aSyn and tau tangles had independent associations with olfactory function, highlighting the importance of using biomarkers of both neuropathologies. Further, longitudinal decline was associated with aSyn, suggesting that incidental Lewy body disease is associated with olfactory decline and reinforcing the known strong association between aSyn and olfactory decline.^37–39^

### Limitations

While cognitive assessments were performed yearly, olfactory testing was only conducted every three years. Time of conversion was defined as the last assessment where an individual was cognitively unimpaired, however, incident MCI may occur between yearly assessments. Nevertheless, diagnosis of MCI/dementia was performed through yearly consensus conferences including the review of full neuropsychological and cognitive clinician evaluation. To ensure the results were not driven by the older age of converter participants, we age-matched groups at the time of conversion, rather than correcting for age in the models. While this approach removed age related biases, it also limited the availability of data points after conversion for the non-converter group. Nonetheless, considering the strong relationship between aging and olfactory decline, age-matching remains a strength of this study and does not limit the assessment of the pre-conversion period, which is of greater significance for evaluating the clinical utility of olfactory testing as an early marker of cognitive decline. Finally, our study population is predominantly white/Caucasian and participants are enrolled as normal aging controls. Our findings may therefore be limited in generalizability, and the potential for volunteer participation bias should be considered.

## Conclusions

We report significantly lower olfactory identification performance up to a decade before conversion to MCI and significant declines with aging over time. This decline was associated with increased neuropsychiatric symptoms, motor impairment, and memory decline, as well as higher burden of neuropathology markers, especially tau NFT and aSyn. These findings support olfactory testing as a potential early clinical marker of neurodegenerative processes preceding MCI.

## Data Availability

The data that support the findings of this study are available from the corresponding author upon reasonable request.

## Author Contributions

Tremblay and Zeighami had full access to all the data in the study and takes responsibility for the integrity of the data and the accuracy of the data analysis.

Study concept and design: Tremblay, Zeighami, Dadar, Beach, Adler

Acquisition, analysis, or interpretation of data: All authors.

Statistical analysis: Zeighami, Tremblay, Dadar

Drafting of the manuscript: Tremblay, Zeighami

Critical revision of the manuscript for important intellectual content: All authors.

